# Factors Associated with Relative Handgrip Strength among Medical Students in Serbia: An Exploratory Cross-Sectional Study

**DOI:** 10.64898/2026.08.24.26361230

**Authors:** Konstantinos Stratakis, Dino Adrović, Zorica Terzić-Šupić, Zorana Nikolov, Jovana Todorović

**Affiliations:** Faculty of Medicine, University of Belgrade, Belgrade, Serbia; Institute of Social Medicine, Faculty of Medicine, University of Belgrade, Belgrade, Serbia

**Author notes:** **Corresponding author: Konstantinos Stratakis**.

**Keywords:** handgrip strength, relative handgrip strength, muscular strength, medical students, physical activity, depressive symptoms, body mass index

## Abstract

Handgrip strength (HGS) is a practical measure of muscular strength, but the factors associated with relative HGS among young adults remain insufficiently understood. This exploratory cross-sectional study examined demographic and anthropometric characteristics, total physical activity (PA), and depressive symptoms in relation to relative HGS among medical students in Serbia. A total of 424 students from the Faculty of Medicine, University of Belgrade, completed HGS testing and study questionnaires. HGS was measured using a VALD DynaMo Lite dynamometer and expressed relative to body weight (N/kg). Participants were categorized according to sample-specific relative HGS percentiles: <25th, 25th–75th, and >75th percentile. PA was assessed using the IPAQ-SF and depressive symptoms using the PHQ-9. Three pairwise multivariable logistic regression models were used to examine factors associated with relative HGS categories. Sex and BMI were the most consistent correlates of relative HGS. Female students had lower odds of belonging to higher relative HGS categories, while higher BMI was consistently associated with lower odds of belonging to higher categories. Age was associated only with the 25th–75th percentile group compared with the <25th percentile group. Total PA volume and PHQ-9 score were not independently associated with relative HGS categories. These findings suggest that relative HGS in this population was more strongly associated with demographic and anthropometric characteristics than with total self-reported PA or depressive symptom severity.

## Introduction

Muscular strength is a fundamental component of physical fitness that supports everyday function, physical independence, and overall health. Accordingly, lower muscular strength has been associated with adverse health outcomes across the lifespan (McLeod et al., 2016; García-Hermoso et al., 2018). Handgrip strength (HGS) is widely used as a measure of muscular strength because it is quick to assess, requires minimal equipment, and demonstrates good reliability (Reuter et al., 2011; Gränicher et al., 2024). Besides its practicality, HGS has been associated with clinically important outcomes, including all-cause mortality, cardiovascular mortality, and disability. (Soysal et al., 2021).

The way HGS is expressed is important when individuals of different body sizes are compared. Absolute HGS represents total force production but is influenced by body size and muscle mass. Relative HGS, commonly expressed in relation to body weight, provides a strength-to-body-mass measure and therefore offers a different perspective on muscular strength. Absolute and relative HGS should not be considered interchangeable, nor should one be regarded as universally superior to the other, as normalization can influence their relationships with other characteristics (Nevill et al., 2022). In young adults, relative HGS has been associated with lower body fat and a more favorable metabolic profile, whereas absolute HGS appears more strongly related to lean mass (Pettersson-Pablo et al., 2024; Abdelnour et al., 2025). Relative HGS may therefore be particularly informative when comparing individuals of different body sizes and examining strength in relation to other health and lifestyle characteristics.

Several individual characteristics remain important when relative HGS is examined. Sex is one of the most consistent determinants of HGS, with males generally demonstrating higher values than females (Dodds et al., 2014; de Lima et al., 2017). These differences are partly related to skeletal muscle mass, body composition, and upper-limb characteristics. Anthropometric characteristics are similarly relevant. HGS has been associated with fat-free mass, fat mass, overall body size, and upper-limb characteristics (Zaccagni et al., 2020; Myles et al., 2025). In the context of relative HGS, body mass and composition are particularly important because the same absolute force may represent substantially different relative strength in individuals of different body sizes. Age is another established determinant across the lifespan, with strength generally increasing from adolescence into early adulthood before declining later in life (Dodds et al., 2014). Sex, age, and anthropometric characteristics therefore provide an important background against which other potential correlates of relative HGS can be considered.

Beyond these established characteristics, behavioral factors may also contribute to differences in muscular strength. Physical activity (PA) is one potential correlate, although its relationship with HGS is not straightforward. Resistance and muscle-strengthening activities would be expected to relate more directly to strength than walking or predominantly aerobic activity. Studies have reported inconsistent associations between PA and HGS (Tsekoura et al., 2023; Gråstén et al., 2026). This distinction is relevant when PA is assessed with the IPAQ-SF, which captures walking and moderate- and vigorous-intensity activity accumulated across daily life but does not specifically quantify resistance exercise. Consequently, a high total PA score may reflect substantial walking or aerobic activity without necessarily indicating regular strength training. Total PA may therefore relate to relative HGS without directly reflecting strength-specific activity.

Psychological characteristics may also be relevant. Lower HGS has been associated with depressive symptoms (Ren et al., 2020; Cao et al., 2021), while prospective meta-analyses also support an association between lower HGS and subsequent depression (Huang et al., 2021; de Oliveira et al., 2026). Depressive symptoms commonly include low mood, fatigue, anhedonia, sleep disturbance, and reduced motivation, which may limit participation in physical activity and reduce willingness to perform physically demanding tasks (Horne et al., 2021). PA and depressive symptoms are also inversely associated, with more active individuals generally reporting fewer depressive symptoms (Huang et al., 2025; Wang et al., 2026). In addition, inflammatory, neurotrophic, and neuroendocrine pathways have been discussed as possible links between exercise and mental health (da Cunha et al., 2023; Montgomery and Grant, 2026). Because PA and depressive symptoms may influence one another and may also be related to strength, considering both variables together may help clarify their independent associations with relative HGS.

Taken together, relative HGS may be associated with characteristics from several domains. Sex, age, and anthropometric characteristics are relatively established correlates, whereas relationships with total PA and depressive symptoms appear less consistent. Examining these factors together may provide a broader picture of the characteristics independently associated with relative HGS without assuming a single causal pathway. Medical students represent a relevant population in which to explore these relationships. Medical education involves substantial academic demands, while depressive and anxiety symptoms are common across medical-student populations (Agyapong-Opoku et al., 2026). PA has also been associated with well-being and lower burnout in medical students, suggesting that activity patterns may be relevant to both physical and psychological health in this group (Taylor et al., 2022). However, relatively little research has examined relative HGS and its correlates specifically in this population. Accordingly, the present exploratory cross-sectional study aims to investigate whether demographic and anthropometric characteristics, total PA, and depressive symptoms are associated with relative HGS among medical students in Serbia.

## Methods

### Study design and participants

This exploratory cross-sectional study was conducted during the last week of November 2024 among medical students at the Faculty of Medicine, University of Belgrade, Serbia. Data were collected during Social Medicine classes. Students attending classes during the study period were informed about the study aims and procedures and were invited to participate voluntarily. Written informed consent was obtained before data collection.

A total of 431 students were invited to participate. Complete HGS and questionnaire data were available for 424 participants, resulting in a final analytic sample of 424 students and a completion rate of 98.4%. The study was approved by the Ethics Committee of the Faculty of Medicine, University of Belgrade (No. 25/IX-4).

### Measures

Data included sex, age, self-reported body height and body weight, PA, depressive symptoms, and directly measured HGS. BMI was calculated as body weight in kilograms divided by height in meters squared (kg/m^2^).

PA was assessed using the International Physical Activity Questionnaire Short Form (IPAQ-SF) (Hagströmer et al., 2006). The IPAQ-SF assesses walking, moderate-intensity PA, and vigorous-intensity PA performed during the previous seven days. Total PA volume was calculated according to IPAQ-SF scoring procedures and expressed as metabolic equivalent task minutes per week (MET-minutes/week).

Depressive symptoms were assessed using the Patient Health Questionnaire-9 (PHQ-9) (Kroenke et al., 2001). The PHQ-9 contains nine items assessing depressive symptom severity during the previous two weeks. Items are scored from 0 to 3, producing a total score ranging from 0 to 27, with higher scores indicating greater depressive symptom severity.

HGS was assessed isometrically using the VALD DynaMo Lite dynamometer. Participants were seated with both feet flat on the floor. The tested arm was positioned with the elbow flexed at 90°, the upper arm close to the trunk, and the forearm and wrist in a neutral position. Participants were instructed to squeeze the dynamometer with the dominant hand as forcefully as possible for five seconds. Three trials were performed with 30 seconds of rest between trials, and the highest recorded value was used as maximal dominant-hand HGS. To account for differences in body mass, HGS was expressed relative to body weight in Newtons per kilogram (N/kg). Participants were then categorized according to the distribution of relative dominant-hand HGS in the analytic sample: <25th percentile, 25th– 75th percentile, and >75th percentile. These categories represented lower, middle, and higher relative HGS within the study sample.

### Statistical analysis

Descriptive and analytical statistics were used. Categorical variables were presented as frequencies and percentages, while continuous variables were presented as mean ± standard deviation. Normality was assessed using the Kolmogorov-Smirnov and Shapiro-Wilk tests. Differences between relative HGS categories were examined using the chi-square test for categorical variables and the Kruskal-Wallis test for continuous variables without a normal distribution.

Variables showing statistically significant differences (p<0.05) between relative HGS categories in the initial analyses—sex, age, BMI, total PA volume, and PHQ-9 score—were entered into the multivariable models. Relative HGS category was treated as the outcome variable.

Three pairwise binary logistic regression models were constructed: (1) >75th percentile versus <25th percentile, (2) 25th–75th percentile versus <25th percentile, and (3) >75th percentile versus 25th–75th percentile. Results were reported as odds ratios (ORs) with 95% confidence intervals (CIs). All analyses were performed using IBM SPSS Statistics version 22.0.

## Results

The 25th, 50th, and 75th percentiles of relative dominant-hand HGS were 2.71, 3.09, and 3.44 N/kg, respectively. Participants across the relative HGS categories differed significantly by sex (p=0.001), age (p=0.007), BMI (p=0.001), total PA volume (p=0.005), and PHQ-9 score (p=0.022). Participant characteristics according to relative HGS category are presented in Table 1.

**Table 1.** Characteristics of participants according to relative dominant-hand HGS percentile categories.

| Characteristics | <25<sup>th</sup> percentile | 25<sup>th</sup>-75<sup>th</sup> percentile | >75<sup>th</sup> percentile | P-value |
| --- | --- | --- | --- | --- |
| Sex (N%) |  |  |  |  |
| Male | 17 (16.2) | 43 (20.2) | 65 (61.3) |  |
| Female | 88 (83.8) | 170 (79.8) | 41 (38.7) | **0.001** |
| Age in years X±SD | 23.26±1.09 | 23.62±1.24 | 21.63±1.78 | **0.007** |
| BMI in kg/m<sup>2</sup> X±SD | 24.43±3.76 | 22.81±3.46 | 22.55±3.52 | **0.001** |
| Total PA volume in MET-minutes/week X±SD | 2219.13±1580.16 | 2922.04±1930.01 | 2881.49±2102.17 | **0.005** |
| PHQ-9 score X±SD | 9.72±5.96 | 10.71±5.52 | 11.47±6.25 | **0.022** |

*Values are presented as n (%) for categorical variables and mean ± standard deviation for continuous variables. Percentages are calculated within each relative HGS percentile group. Group differences were examined using the chi-square test for categorical variables and the Kruskal-Wallis test for continuous variables. BMI, body mass index; HGS, handgrip strength; MET, metabolic equivalent task; PA, physical activity; PHQ-9, Patient Health Questionnaire-9.*

In the adjusted model comparing the >75th percentile with the <25th percentile, female sex was associated with lower odds of belonging to the >75th percentile group (OR=0.01, 95% CI 0.003–0.04), as was higher BMI (OR=0.59, 95% CI 0.49–0.68). In the comparison between the 25th–75th and <25th percentile groups, female sex was associated with lower odds of belonging to the 25th–75th percentile group (OR=0.27, 95% CI 0.12–0.61), while older age was associated with higher odds (OR=1.38, 95% CI 1.08–1.77). Higher BMI was again associated with lower odds of belonging to the 25th–75th percentile group (OR=0.81, 95% CI 0.74–0.88).

Finally, in the model comparing the >75th percentile with the 25th–75th percentile group, female sex (OR=0.03, 95% CI 0.01–0.80) and higher BMI (OR=0.73, 95% CI 0.64–0.82) were associated with lower odds of belonging to the >75th percentile group. Total PA volume and PHQ-9 score were not independently associated with relative HGS category in any of the adjusted comparisons. Full multivariable results are presented in Table 2.

**Table 2.** Pairwise multivariable logistic regression analyses of factors associated with relative HGS percentile categories.

| Characteristics | >75<sup>th</sup> percentile vs.<br><25<sup>th</sup> percentile | 25<sup>th</sup>-75<sup>th</sup> percentile vs.<br><25<sup>th</sup> percentile | >75<sup>th</sup> percentile vs.<br>25<sup>th</sup>-75<sup>th</sup> percentile |
| --- | --- | --- | --- |
| Sex |  |  |  |
| Male | 1.0 | 1.0 | 1.0 |
| Female | **0.01 (0.003-0.04)** | **0.27 (0.12-0.61)** | **0.03 (0.01-0.8)** |
| Age in years | 0.98 (0.94-1.02) | **1.38 (1.08-1.77)** | 0.97 (0.93-1.02) |
| BMI in kg/m<sup>2</sup> | **0.59 (0.49-0.68)** | **0.81 (0.74-0.88)** | **0.73 (0.64-0.82)** |
| Total PA volume in MET-minutes/week | 1.00 (1.00-1.00) | 1.00 (1.00-1.00) | 1.00 (1.00-1.00) |
| PHQ-9 score | 1.04 (0.98-1.11) | 1.03 (0.99-1.08) | 1.01 (0.96-1.02) |

*Values are presented as odds ratios with 95% confidence intervals. Relative HGS percentile categories were defined according to dominant-hand HGS expressed relative to body weight in N/kg. The <25th percentile group was used as the reference category in the first two models, and the 25th–75th percentile group was used as the reference category in the third model. Male sex was used as the reference category. BMI, body mass index; HGS, handgrip strength; MET, metabolic equivalent task; PA, physical activity; PHQ-9, Patient Health Questionnaire-9.*

## Discussion

This exploratory cross-sectional study examined factors associated with relative HGS among medical students. Sex and BMI were the most consistent correlates. Age showed an isolated association, whereas total self-reported PA and PHQ-9 score were not independently associated with relative HGS. Overall, relative HGS appeared more closely related to demographic and anthropometric characteristics than to total PA or depressive symptoms. Female students had lower odds of belonging to higher relative HGS categories than male students. This is consistent with extensive evidence showing higher HGS among males across adult populations (Dodds et al., 2014; de Lima et al., 2017; Steiber, 2016). Differences in skeletal muscle mass, body composition, and upper-limb characteristics may contribute to this pattern (Janssen et al., 2000; Zaccagni et al., 2020; Myles et al., 2025). In this sample, the sex association remained after adjustment for BMI, although BMI cannot capture differences in fat-free mass or regional muscle distribution.

BMI was consistently inversely associated with relative HGS. Similar patterns have been reported in young adults, with relative HGS showing strong inverse relationships with body fat and other adiposity-related characteristics (Pettersson-Pablo et al., 2024; Abdelnour et al., 2025). Obesity and higher adiposity may also influence relative muscle function through changes in mobility, muscle morphology, and metabolic or inflammatory factors (Tomlinson et al., 2016; Pettersson-Pablo et al., 2024). However, this finding should be interpreted cautiously. BMI does not distinguish fat from lean tissue, and body weight was included directly in the relative HGS calculation. Future studies using direct body-composition measures could clarify whether adiposity, muscle mass, or the mathematical construction of relative HGS primarily explains this result.

Age showed a less consistent association. Older age was associated with higher odds of being in the 25th–75th percentile group rather than the <25th percentile group, but not with the other comparisons. Given the relatively narrow age distribution in the sample, the ability to detect a clear age-related pattern may have been limited. This isolated finding may reflect sampling variability or unmeasured differences in training and lifestyle and should therefore be interpreted cautiously.

Total PA volume was not independently associated with relative HGS. General PA includes activities that provide different stimuli for strength development. Resistance training and progressive high-force exercise are more directly related to strength development than walking or many forms of aerobic activity (American College of Sports Medicine, 2009; Garber et al., 2011; Suchomel et al., 2018). The IPAQ-SF does not specifically assess resistance training, exercise progression, or training volume. Consequently, high total MET-minutes/week may not indicate substantial strength-focused activity. This may explain the mixed findings reported in student populations (Tsekoura et al., 2023; Gråstén et al., 2026). The absence of an association does not mean that PA is irrelevant to muscular strength. Rather, total self-reported PA may be too nonspecific to capture activities most relevant to HGS. Future studies should distinguish resistance, aerobic, occupational, and recreational activity and should include objective measures where possible.

The difference in PHQ-9 score observed across HGS categories in the initial analysis was not evident in the adjusted models that included sex, age, BMI, and PA. This differs from studies reporting inverse adjusted associations between HGS and depressive symptoms (Ren et al., 2020; Cao et al., 2021; Huang et al., 2021; de Oliveira et al., 2026). Differences in HGS normalization, depression measures, populations, and covariate adjustment may explain the discrepancy. Depressive symptoms and physical fitness may also be linked indirectly through fatigue, sleep disturbance, motivation, and activity behavior (Horne et al., 2021). Thus, these findings indicate only that PHQ-9 score was not independently associated with relative HGS in this sample.

The study has several strengths, including direct HGS measurement using a standardized dynamometer protocol, a high completion rate, and a focus on a relatively understudied young medical-student population. However, the cross-sectional design prevents causal inference. The single-faculty sample limits generalizability, and self-reported height and weight may have affected both BMI and relative HGS. BMI also does not reflect body composition. PA was assessed using the IPAQ-SF, which is subject to recall bias and does not provide detailed information on resistance exercise. Finally, sample-specific HGS percentiles limit comparisons with external populations, and the use of three pairwise models without adjustment for multiple comparisons means that isolated findings, particularly the age association, should be interpreted cautiously.

## Conclusion

Among medical students, relative HGS was most consistently associated with sex and BMI. Female students had lower odds of belonging to higher relative HGS categories, while higher BMI was associated with a lower strength-to-body-mass ratio. Age showed an inconsistent association, and total self-reported PA and PHQ-9 score were not independently associated with relative HGS.

These findings suggest that demographic and anthropometric characteristics were more consistently associated with relative HGS than overall PA volume or depressive symptom severity in this population. However, the BMI finding should not be interpreted as evidence of lower absolute strength. Longitudinal studies using objective body-composition measures and detailed assessments of resistance exercise are needed to clarify the determinants and relevance of relative HGS in young adults.

## Contributions

K. S. contributed to the conceptualization, design, data gathering, data analysis, writing, and correcting the article. D. A. contributed to the conceptualization, design, data gathering, data analysis, writing, and correcting the article. Z. T. contributed to the conceptualization, design, writing, and correcting of the article and provided supervision. Z. N. contributed to the conceptualization, data gathering, writing, and correcting of the article. J. T. contributed to conceptualization, design, data gathering, data analysis, writing, and correcting the article and provided supervision.

## Funding

This research received no specific grant from any funding agency in the public, commercial, or not-for-profit sectors.

## Data availability statement

The datasets used and/or analyzed during the current study are available from the corresponding author upon reasonable request.

## Acknowledgments

The authors thank the participants who made the study possible.

## Conflicts of interest

The authors declare no conflicts of interest.

## Notes

### Competing Interest Statement

The authors have declared no competing interest.

### Author Declarations

The study was approved by the Ethics Committee of the Faculty of Medicine, University of Belgrade (No. 25/IX-4).

